# Evaluation of Urinary Circulating Tumor DNA to Detect Minimal Residual Disease in Patients with High-Risk Non-Muscle Invasive Bladder Cancer

**DOI:** 10.64898/2026.09.09.26362655

**Authors:** Roger Li, Joshua Linscott, Prithvi Murthy, Kyle Rose, Young Pak, Frank Zhang, Yong Huang, G. Daniel Grass, Jonathan Chatzkel, Noah M. Hahn, David J. McConkey, Joshua J. Meeks, Siamak Daneshmand, Thomas W. Flaig, Seth P. Lerner, Logan Zemp, Michael A. Poch, Philippe E. Spiess, Wade J. Sexton, Scott M. Gilbert, Hongzhi Xu, Pan Du, Shidong Jia, Xuefeng Wang

## Abstract

**Purpose:** Restaging transurethral resection of bladder tumor (reTURBT) is the standard-of-care for some patients with high-risk non-muscle invasive bladder cancer (NMIBC). However, it may lead to complications and biomarkers to avoid unnecessary reTURBT are needed.

**Materials and Methods:** Patients with high-risk NMIBC undergoing reTURBT were prospectively enrolled. Whole-exome sequencing along with low-pass whole-genome sequencing were performed on index TURBT samples to generate mutational profiles and copy number variation. Up to 50 personalized mutations and a fixed panel of 500 hotspot mutations were used for detecting variants from urine samples collected prior to reTURBT. Using a prespecified algorithm, tumor fraction (TF) and copy number burden (CNB) score were used to measure minimal residual disease and correlated with reTURBT pathology.

**Results:** Overall, 72/76 urine samples collected from patients with high-risk NMIBC prior to reTURBT passed quality check. From the indext TURBT specimens, a median of 39 personalized variants from the index TURBT were used for disease tracking. Overall, median TF was 0.206 vs. 0.001 in patients with and without residual tumor, respectively (p<0.001). Similarly, CNB score was elevated (7.34 vs. 4.02, p<0.001). utDNA achieved sensitivity of 97.8% and specificity of 69.2%, with area under the curve (AUC) of 0.932 in predicting disease found on reTURBT. Analyses of the index and reTURBT tissue and urine mutational profiles provided evidence for underdiagnosis of subclinical disease at the time of reTURBT.

**Conclusion:** Paired personalized and panel utDNA can be used to accurately detect residual disease in patients with high-risk NMIBC with potentially wide-ranging clinical applications.

## Introduction

Staging of non-muscle invasive bladder cancer (NMIBC) is achieved using transurethral resection of bladder tumor (TURBT). However, quality of TURBT varies, leaving significant risk for understaging and disease persistence, with most of the residual lesions found at the original tumor location.^1^ As such, repeat TURBT (reTURBT) is often performed for patients with high-risk NMIBC^2–4^, leading to improved oncologic outcomes.^5^ However, TURBT is known to cause deficits in general and mental health perception as well as decreased physical and social functioning.^6^ Clinically significant complication rates occur in 5-6%, including significant bleeding (2.3-2.8%) and bladder perforation (1.3-3.5%).^7,8^ Biomarkers to identify patients with residual disease following primary TURBT are thus critically needed.

Bladder cancer is unique in that malignant cells are constantly interfacing the urine. For decades, researchers attempted to leverage this easy-to-access body fluid to detect cancer.^9^ The advent of next-generation sequencing (NGS) has allowed comprehensive molecular analyses of tumor and liquid biopsy samples. Recently, several studies have explored the use of urinary circulating tumor DNA (utDNA) for disease detection and monitoring.^10–12^ In this study, we used a combined personalized and paneled utDNA approach to understand whether residual disease following index TURBT (iTURBT) can be prospectively identified using urine samples collected prior to standard-of-care (SOC) reTURBT.

## Material and Methods

### Patient Selection and Sample Collection

Following Institutional Review Board approval (MCC21616) and informed consent, patients with diagnosis of HG NMIBC undergoing reTURBT at the discretion of the treating physician were prospectively enrolled in the discovery cohort from December 2021 to July 2023. Pathologic specimens were reviewed by board-certified genitourinary pathology specialists and classified according to the AJCC Cancer Staging System.^13^ reTURBT was performed between 6-18 weeks from initial diagnosis, accounting for delays in referral, using white light or photodynamic diagnosis (PDD) according to treating physician discretion. Urine (preUR) (40mL) and matched PBMC samples were collected immediately prior to reTURBT (**Supplemental Fig. 1**).

### Minimal Residual Disease Assay Design

The primary objective was to investigate the ability of utDNA to identify patients with minimal residual disease (MRD) as corroborated on reTURBT. Recapitulating the clinical workflow, whole-exome sequencing (WES) of the tumor samples obtained from iTURBT and patient PBMC specimens was performed to detect and censor germline and CHIP mutations. Variants were detected with variant allele frequencies (VAF) down to 0.1% (novel mutations) and 0.01% (known baseline mutations) as the reference standard to perform the PredicineBEACON personalized MRD assay^14,15^ on the preUR samples. In parallel, low-pass whole genome sequencing (LP-WGS) assay was performed at 3x sequencing depth as previously described.^16^ To verify concordance between genomic alterations found in the iTURBT and reTURBT tumor samples, WES was also performed using reTURBT tumor/benign specimens. Ultra-deep sequencing of preUR was conducted using a personalized panel consisting of somatic mutations identified within the iTURBT tumor specimen and an additional fixed panel covering >500 actionable mutations.

### MRD call

utDNA was extracted from the supernatant of the preUR samples as previously described.^15^ In brief, cfDNA was isolated using an automated magnetic bead–based extraction platform. After quantification and normalization, cfDNA underwent library preparation, which includes end-repair dA-tailing and adapter ligation. Ligated library fragments with appropriate adapters are amplified via PCR. The amplified DNA libraries are then further checked using Agilent TapeStation (Agilent Technologies, CA) and samples with sufficient yield are moved to hybrid capture. To detect a personalized variant selected for MRD tracking, at least one mutant fragment with confident variant support was required. Hotspot mutations not in the personalized sample are considered as novel variants if they have two or more double-stranded mutant fragments. To call a sample as analytical MRD positive, it must contain two or more personalized or novel variants, with one of them found to have double-stranded mutant fragment support.^15^

The tumor fraction (TF) was estimated according to the somatic mutations detected from the MRD assay. TF was estimated on the basis of the mutant and total fragment counts of somatic mutations: 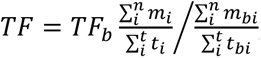, where *TF_b_* is the tumor fraction of the matched baseline sample, *i* is the *ith* selected mutation for MRD tracking, *n* is the total number of selected mutations for MRD tracking, *m* is the number of mutant fragments carrying the mutation, and *t* is the total number of fragments (including wild-type fragments) at the mutation site; *m_b_* and *t_b_* are mutant fragment and total fragment counts at the mutation site of the baseline sample.

Companion LP-WGS with an average coverage of 3x was performed for each sample. The ichorCNA package^17^ was applied for GC and mappability-normalization and to estimate copy number variations (CNV) using the hidden Markov model. A sample-level copy number burden (CNB) score was estimated based on the absolute z-score of the arm-level CNV deviations. CNV abnormality was called based on comparison of the CNB score against scores from a pool of healthy donor urine samples (CNB score ≥ 5.8)^16^. The final MRD tracking was called positive when TF ≥ 0.20% or CNB score was positive; and negative when both TF < 0.20% and CNB score was negative.

### Statistical Analysis

R version 4.3.0 (https://www.R-project.org) was used for statistical analysis and graphic plotting with ggplot2 (RRID:SCR_ 014601) and ComplexHeatmap (RRID:SCR_017270) packages. The R package pROC was used to perform ROC analysis. The copy number G-score was calculated by the GISTIC (RRID:SCR_000151) 2.0 pipeline via Gene-Pattern (RRID:SCR_003201, https://www.broadinstitute.org/cancer/ software/genepattern). The Wilcoxon rank-sum or Student t test was used to compare numeric variables. Fisher exact test was used to compare categorical variables. All tests were two sided and considered statistically significant at p<0.05.

## Results

### Patient Characteristics and Outcomes

A total of 76 patients with HG NMIBC undergoing reTURBT at Moffitt Cancer Center were enrolled. Baseline clinicopathologic characteristics are displayed in **Table 1**. Of these patients, 49 (64.5%) were staged T1, 20 (26.3%) Ta, and 7 (9.2%) Tis. Overall, 29 (38.2%) had recurrent NMIBC and 19 (25%) having previously received intravesical treatment. Mixed histologic subtype was found either in the index or repeat TURBT sample in 10 (13.2%) patients. PDD was used at the time of reTURBT in 11 patients (14.5%) according to surgeon discretion. On repeat TURBT, 50 patients (65.8%) were found to have residual tumor, with 4 (5.3%) upstaged to MIBC, 19 (25.0%) with T1HG, 15 (19.7%) TaHG, 11 (14.5%) Tis and 1 (1.3%) TaLG. Following DNA extraction, 4 failed QC, and 72 preUR samples were used for analysis.

**Table 1.** Clinicopathologic characteristics of patients with high-risk non-muscle invasive bladder cancer in the discovery and validation cohorts.

| Characteristic | Discovery<br>N = 76<sup>1</sup> |
| --- | --- |
| **Sex** |  |
| Male | 60 (78.9%) |
| **Race** |  |
| White | 71 (93.4%) |
| Other | 5 (6.6%) |
| **ECOG** |  |
| 0 | 58 (76.3%) |
| 1 | 16 (21.2%) |
| 2 | 2 (2.6%) |
| **Prior NMIBC** |  |
| None | 47 (61.8%) |
| Recurrent | 29 (38.2%) |
| **Prior Intravesical Therapy** |  |
| No Prior Therapy | 19 (25.0%) |
| Prior Therapy | 10 (13.2%) |
| **Index TURBT Tumor Stage** |  |
| Tis | 7 (9.2%) |
| TaHG | 20 (26.3%) |
| T1 | 49 (64.5%) |
| ≥T2 | 0 (0.0%) |
| **CIS Index or Repeat TURBT** | 14 (18.4%) |
| **Variant Subtype** | 10 (13.2%) |
| **TURBT Lymphovascular Invasion** |  |
| Absent | 72 (94.7%) |
| Present | 4 (5.3%) |
| **BlueLight at ReTURBT** | 11 (14.5%) |

<sup>1</sup>n (&); Median (Q1, Q3)

### Mutational Landscapes of the Index and Repeat Tissue Samples

In theory, utDNA from the preUR specimen reflects the mutational landscape of the residual tumors present at reTURBT. To justify using iTURBT samples as the reference standard to inform the personalized PredicineBEACON assay, we examined the concordance between the mutational landscapes of the iTURBT and reTURBT specimens (**Fig. 1**). Higher numbers of alterations were found from the iTURBT (n=76, median 117, range 2-1,334) than the reTURBT specimens (n=75, median 14.5, range 0-1,025). Overall, 26.3% of all detected iTURBT alterations were found in the reTURBT specimens. A median of 39 alterations (range 4-50) per patient were used for personalized MRD testing. Importantly, 57.9% (IQR 17.7-87.3%) of alterations observed in iTURBT were found in malignant reTURBT samples, but only 1.2% (IQR 0.4-4.1%) were found with benign tissues. Conversely, >80% of the alterations found in the reTURBT specimens were present in iTURBT samples. Taken together, there appears to be sufficient overlap between the mutational landscape within the iTURBT and reTURBT specimens to support using iTURBT specimen as the reference standard for MRD detection (**Supplemental Fig. 1**).

**Figure 1.**
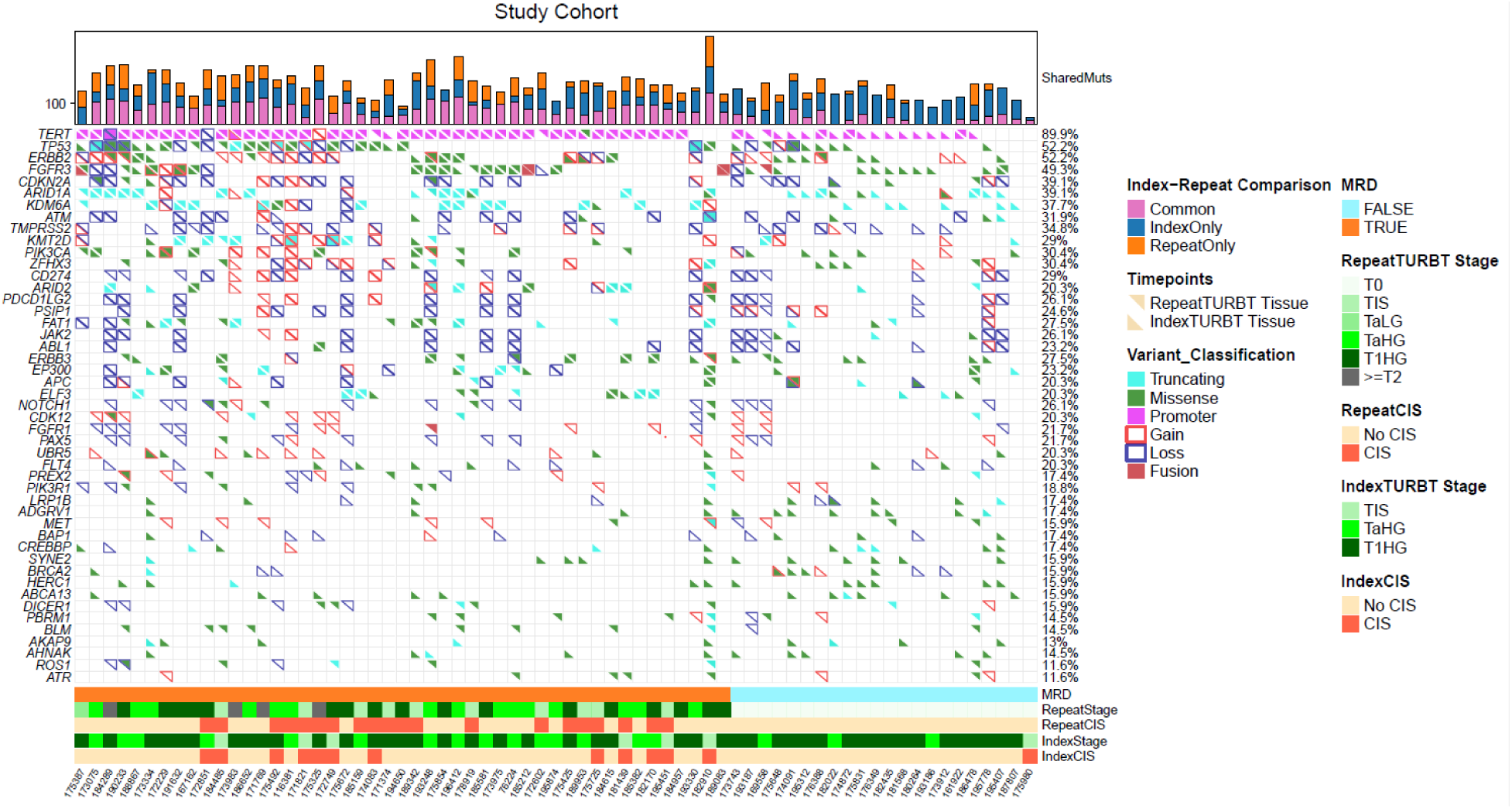
Mutational landscape of index and repeat transurethral resection of bladder tumor (reTURBT) samples. Oncoprint depicting the most frequently observed genomic variants from the index TURBT specimen (lower left triangles) and overall mutational frequencies. Matched and unmatched mutations identified in reTURBT specimens are shown as upper right triangles. Top bars represent total variants detected per patient in index (blue) and reTURBT (orange) specimens, with number of shared variants indicated in purple. Bottom annotations include pathologic minimal residual disease (MRD) status, T stage, and presence of CIS for each patient

Field cancerization is a well-described phenomenon whereby histologically benign urothelial cells within the bladder acquire pro-tumorigenic mutations shared with tumor cells.^18,19^ To rule out the possibility that utDNA can originate from these benign cells thus obfuscating the assay results, we performed WES on the histologically benign tissue samples obtained at reTURBT to understand the prevalence of intrinsic genomic alterations and their VAFs. Of all detected variants in iTURBT samples, 5.1% were also found in the corresponding benign reTURBT specimens with mean VAF of 11.6% (median 1.0%, ranging from 0 to 48.8%, **Fig. 2**). Compared to patients with MRD at reTURBT, the mutational concordance rate was much lower in patients found to have benign reTURBT (**Fig. 2**). Similarly, among all alterations identified in reTURBT tissue that overlapped with the fixed hotspot panel, only 4.2% originated from the benign reTURBT tissue. Importantly, none of the personalized nor panel gene variants in the benign reTURBT samples were found above the prespecified MRD level in the matching preUR specimen. In summary, although genomic alterations were detected within benign biopsies, corresponding urinary levels remained below the prespecified MRD threshold, thereby obviating the risk of interfering with assay accuracy.

**Figure 2.**
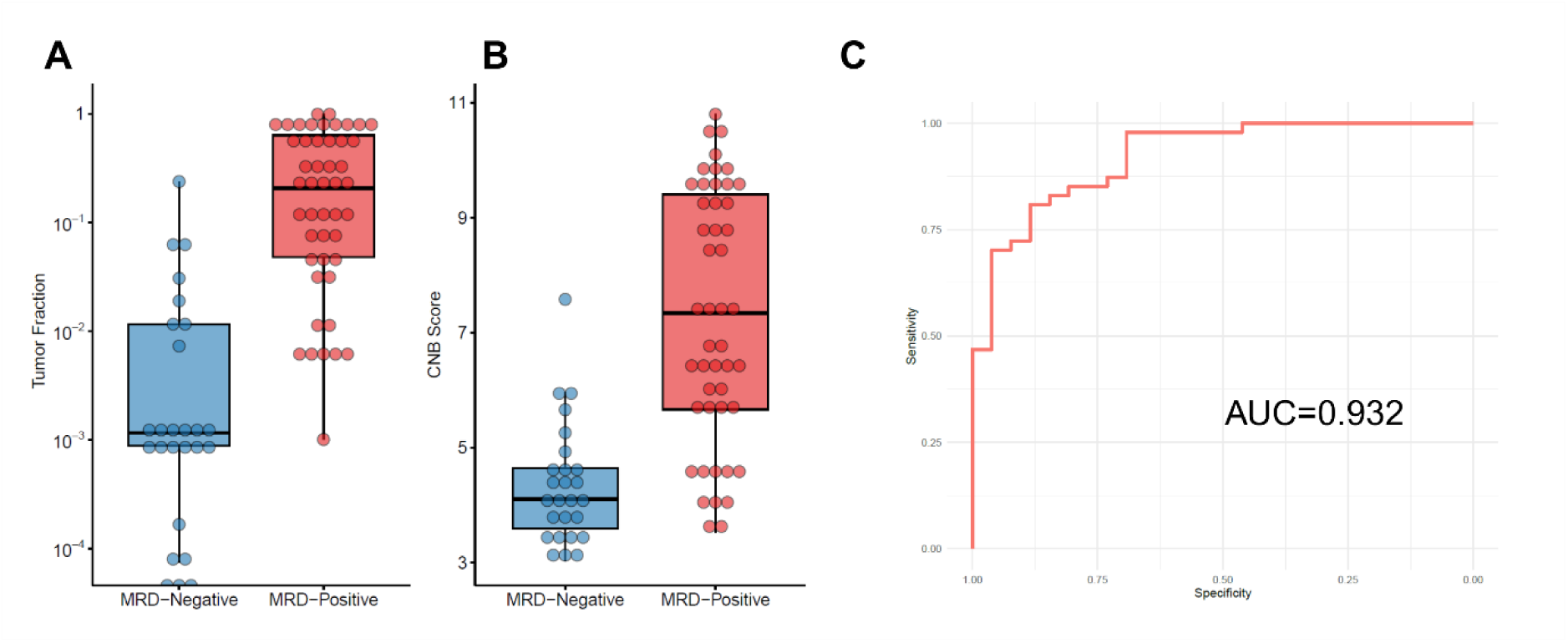
Diagnostic characteristics of tumor fraction (TF) and copy number burden (CNB) score to distinguish between patients with or without minimal residual disease (MRD) at the time of repeat transurethral resection of bladder tumor. a) Urine derived median TF was estimated to be 0.20 (MRD+) vs. 0.0012 (MRD negative-), p<0.001. b) CNB Score for the same groups was estimated at 7.34 vs. 4.02, p<0.001. c) Combining TF at a threshold of TF ≥ 0.002 and CNB Score ≥ 5.8 yielded an area under the receiver operating characteristic curve (AUROC) of 93.2%.

### Detection of utDNA Predicts Minimal Residual Disease

The primary objective of our study was to assess the ability of utDNA to identify patients with MRD as corroborated on reTURBT. Using utDNA extracted from preUR supernatant, TF and CNB score were estimated and used to correlate with reTURBT pathology. Overall, median urine-derived TF was estimated to be 0.206 in patients with residual tumor vs 0.001 in patients with benign reTURBT (p<0.001) (**Fig. 2a**). Similarly, CNB score was found to be elevated in MRD positive patients (7.34 vs. 4.02, p<0.001) (**Fig. 2b**).

Using both patient level TF and CNB Score, utDNA separated the patients into three distinct groups (**Fig. 3**). Those with concordant low TF/CNB were consistently MRD negative (18/19), while others with high TF/CNB were MRD positive (32/33). However, 21 patients with high TF and low CNB demonstrated mixed results, with 14 found to harbor MRD. Using the prespecified MRD threshold of TF ≥ 0.002 and CNB ≥ 5.8, the assay combining TF calculated from personalized mutations and novel mutations in the Fixed Panel, with CNB score achieved sensitivity of 97.8% and specificity of 69.2% at detecting clinical MRD, with an AUC of 0.932 (**Fig. 2c**). This reflects the complementary information provided by CNB scores to the fixed panel and personalized mutations. To further evaluate the contribution of test accuracy gained from the personalized vs. fixed panel testing and CNB scores, we assessed sensitivity and specificities from each independent assay as reported in **Supplemental Fig. 2**.

**Figure 3.**
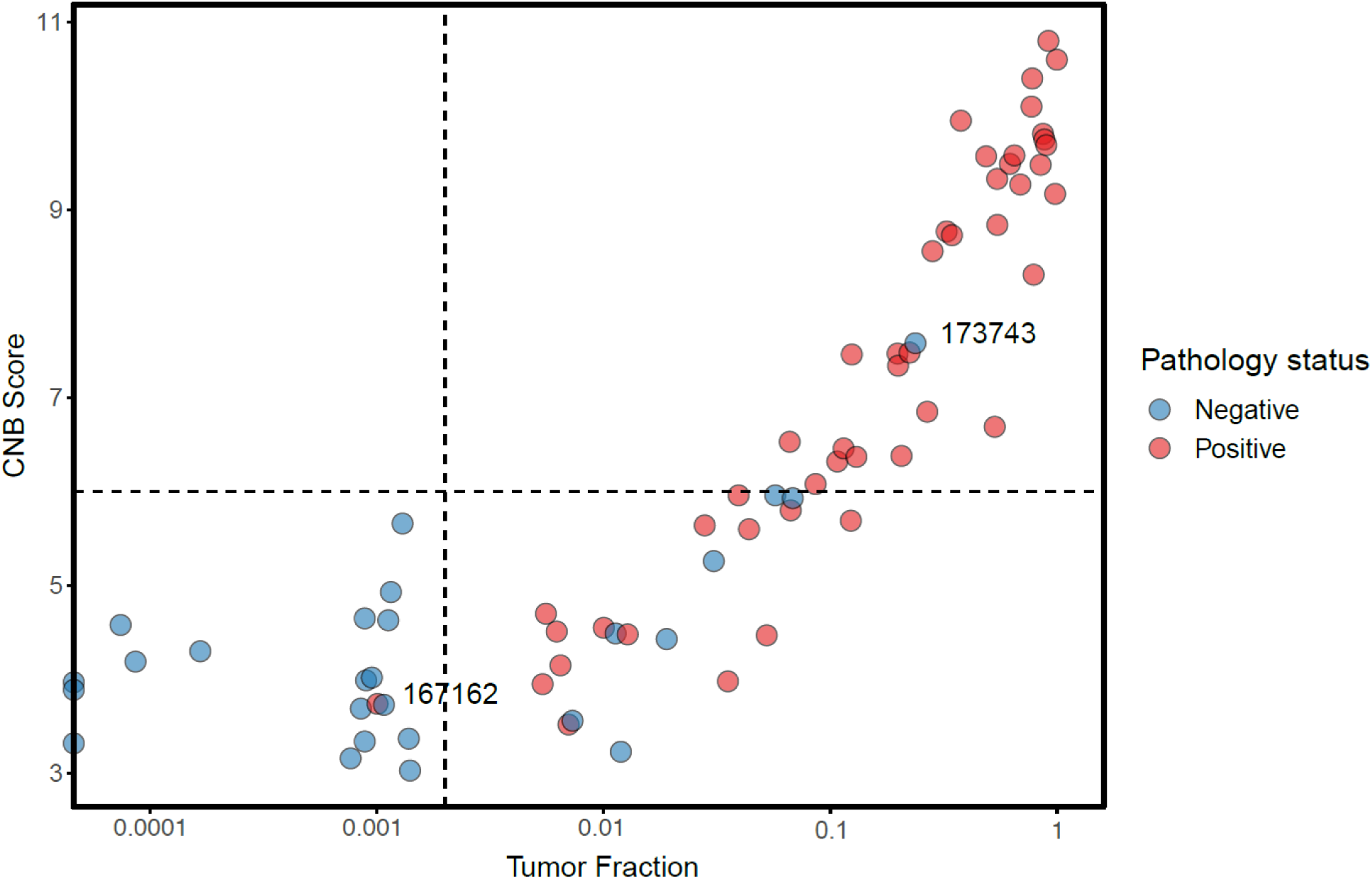
Scatter plot of urinary tumor DNA (utDNA) assay by tumor fraction (TF) and copy number burden (CNB) Score. The utDNA assay performed on urine samples prior to repeat transurethral resection of bladder tumor (reTURBT) stratified patients into three groups. Most patients had concordant utDNA assay and pathology results, however a few discordant particular in those with high TF and low CNB Score were observed. Dashed lines represent predefined thresholds for TF and CNB Score to predict minimal residual disease (MRD). Two patients with outlier utDNA assay versus pathology results are highlighted.

CIS is often multifocal and clinically challenging to identify on cystoscopy.^3^ To evaluate whether the presence of CIS influenced utDNA results, we compared TF and CNB between papillary-only vs. CIS-containing patients (on index or repeat TURBT). Overall, both TF (0.36 vs. 0.18, p<0.01) and CNB (7.42 vs 5.80, p<0.01) were higher in CIS-containing utDNA samples, but the AUCs of MRD detection (0.948 vs 0.949) were not significantly different (p=0.531) between CIS-containing and papillary only tumors.

### Analysis of Discordant Results

To resolve the difference between the utDNA and reTURBT results, we interrogated the gene-level results leading to the discrepant readouts (**Supplemental Table 1**). In the entire study, the only false negative urine assay (167162) detected low levels of personalized *PIK3CA* mutation that was shared between the iTURBT and reTURBT tumors. Interestingly, low levels of a novel *TP53* mutation from the reTURBT specimen were detected via the fixed gene panel. In all of the false positive urine assays, mutations in the iTURBT samples were detected, suggesting subclinical persistence of disease that remained unresected at the time of reTURBT.

Next, we investigated longitudinal follow-up data in patients with available urine samples. In pt 173743, despite markedly elevated TF and CNB, reTURBT was interpreted as benign (**Fig. 3**, **Fig 4A**). Following reTURBT, the patient underwent induction BCG and was found to have T1HG recurrence at first surveillance, strongly suggesting persistence of clinically occult disease during reTURBT. Similarly, utDNA increase was found to predate clinical recurrence in two additional patients (176388, 195312) (**Fig. 4B, C**). In contrast, 2 others (174872, 195778) were found to have prolonged utDNA clearance following induction BCG, and remain disease free at 34.3 and 16.4mo of follow-up, respectively (**Fig 5D, E)**. In summary, results from the longitudinal follow-up data suggest that dynamic changes in utDNA offer promise for disease monitoring and may reflect presence of occult disease.

**Figure 4.**
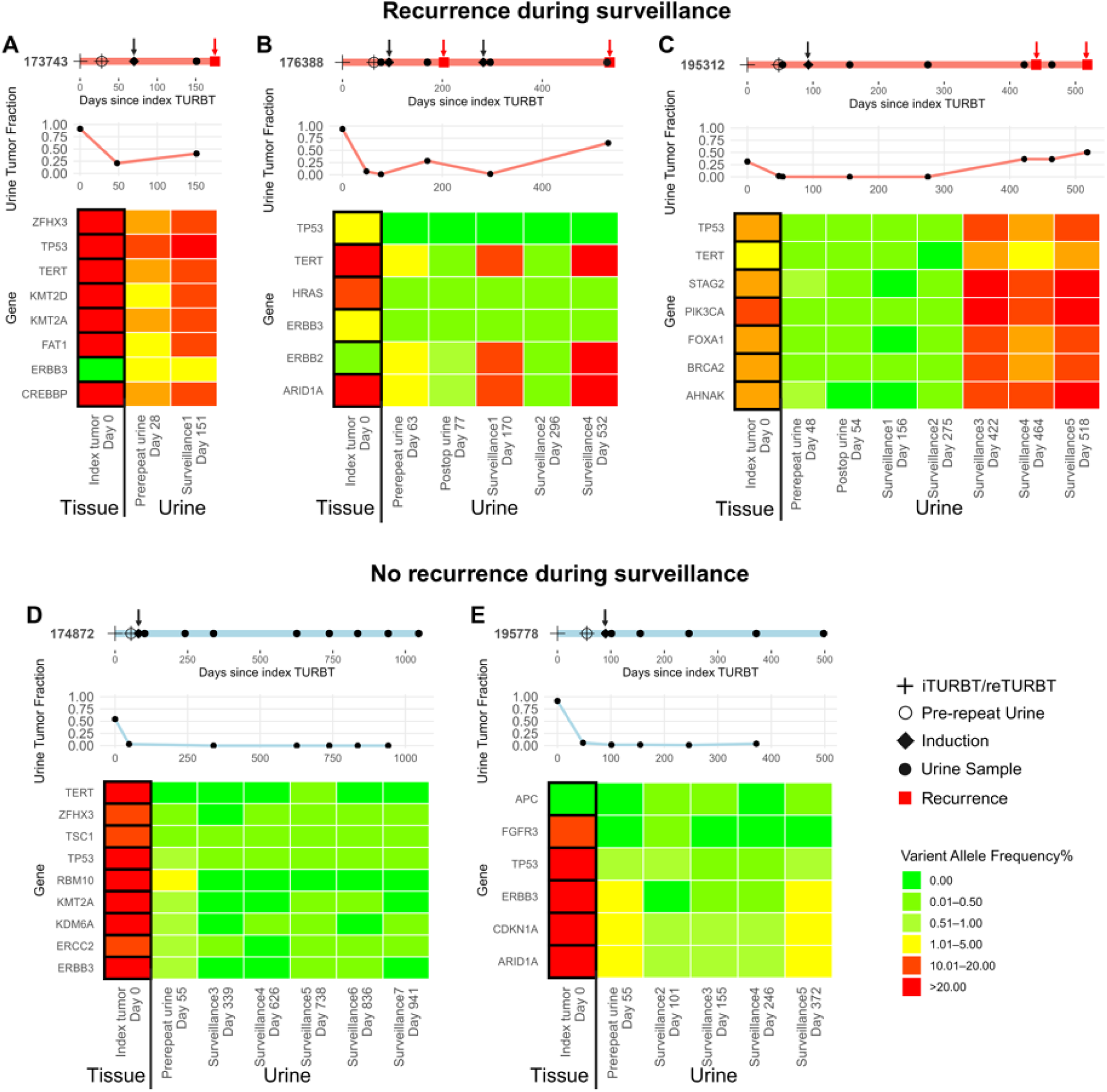
Longitudinal follow-up of urinary tumor DNA (utDNA) in patients with false positive utDNA at the time of repeat transurethral resection of bladder tumor (reTURBT). Top to bottom: (1) swimmer’s plot illustrating key clinical events (legend), with black arrows highlighting intravesical induction and red arrows emphasizing pathologic tumor recurrence; (2) line plot showing total tumor fraction (TF) across index tumor and serial urine samples over time; (3) Heat map displaying variant allele frequencies (VAF) of patient-specific pathologic mutations, comparing the index tumor to follow-up urine, with time from index tumor (in days) labeled on the x-axis.

## Discussion

Using a combined personalized and paneled utDNA assay along with CNB, AUC of 0.932 was achieved to predict the presence of residual disease on reTURBT in patients with high-risk NMIBC. Although genomic variants were found from benign reTURBT samples, illustrating field cancerization, setting the TF threshold ≥ 0.002 for MRD detection was sufficient to distinguish signal (i.e. residual tumor) from noise (field defect). Close examination of the urinary mutational profile and follow-up data from patients found to have false-positive utDNA at the time of testing suggests the presence of occult subclinical disease missed by reTURBT.

Our study demonstrates that combined personalized and panel utDNA enables accurate non-invasive detection of residual disease prior to SOC reTURBT. With a negative predictive value of 95%, this personalized genomic assay can be used to safely avoid unnecessary restaging TURBT in the appropriate patients. When applied to the combined discovery and validation cohorts, reTURBT could have been avoided in 22 patients (26%), while missing a single residual tumor. Despite seemingly lower positive predictive value, utDNA may more accurately reflect the “ground truth” than reTURBT histology, as up to 20% of tumors can be missed by cystoscopic evaluation.^20^ Our in-depth analysis of the utDNA vs. TURBT mutational profiles and clinical follow-up provide additional evidence that utDNA may be tracking disease burden more closely than reTURBT. Additionally, CIS-containing tumors were found to yield higher utDNA readouts, suggesting subclinical disease burden not uncovered at the time of diagnosis. Ultimately, whether quantification of disease burden following diagnostic TURBT can lead to clinically meaningful therapeutic outcomes remains untested.

Beyond its clinical implications, our study additionally yielded several key molecular insights on the genomic landscape of HR NMIBC. Exomic alterations found in our study largely correlated with those previously described.^21,22^ In addition, iTURBT and reTURBT specimens shared sufficient overlap in genomic variants, making it feasible for using WES data from the iTURBT samples as reference to detect utDNA prior to reTURBT. As expected, both VAF from the field cancerization and its corresponding utDNA TF were much lower than those found from carcinomatous samples, making it possible to quantitatively distinguish tumor from normal.

Moreover, the strategy to test both personalized and panel genomic variants not only increases diagnostic specificity, but also enables detection of *de novo* variants arising following treatment.^23^ As demonstrated by the in-depth genomic analysis of the tissue and urine samples from the patient yielding false-negative utDNA results, test accuracy may be improved by lowering the threshold for MRD calling for certain key genomic alterations (e.g. *TP53*, *PIK3CA*). Although less sensitive, CNB may serve as a quantitative measure of clinically detectable disease, indicating the need for cystoscopic intervention. Ultimately, improvements upon the current model will require further testing and validation in larger patient cohorts.

Our study is not without limitations. First, the study cohort is relatively small and single-institutional, exposing our results and interpretation to potential bias and non-generalizability. Future studies leveraging samples collected from multiple institutions are needed to validate the results reported herein. Additionally, incomplete iTURBT or delayed reTURBT was performed on several patients within our cohort, allowing for potential tumor regrowth, increasing the detectability of utDNA. However, dichotomizing our cohort to patients undergoing reTURBT within 6 wks (AUC 0.947) vs. those beyond (AUC 0.939), no difference in AUC was found (p=0.372). Thirdly, not all patients in our cohort underwent reTURBT with enhanced cystoscopy, leaving the possibility of missed occult disease that may have otherwise been detected. Notwithstanding, the clinical conditions seen in patients within this cohort represent real-world reTURBT practice and can readily be applied to a variety of clinical settings.

## Conclusion

Paired personalized and panel utDNA can be used to accurately detect residual disease in patients with HR NMIBC at the time of SOC reTURBT. We show that utDNA as a promising tool for the use of detecting residual disease in the reTURBT setting, with potential wide-ranging applications in several settings in the management of NMIBC.

## Disclosures

**RL**: Research support - Predicine, Veracyte, Johnson & Johnson; Consultant - BMS, Merck, CG Oncology, ImmunityBio, Pfizer, Johnson & Johnson, AstraZeneca, enGene, Valar Labs, Photocure; Travel - Predicine, CG Oncology, Johnson & Johnson; Honoraria - UroToday, IBCG, MashUP Media, MJH Lifesciences

**NMH**: Research support - Predicine, AstraZeneca, Bristol Myers-Squibb, Genentech, Seattle Genetics, Loxo-Oncology, and Incyte; Consultant - AstraZeneca, Merck, Genentech, Ferring, Champions Oncology, Health Advances, Keyquest Health, Guidepoint Global, Seattle Genetics, Incyte, CicloMed, Janssen, Pfizer, Boehringer Ingelheim, EMD Serono, Protara, Verity Pharmaceuticals, and Astellas Pharma; Speaking honorarium - Creative Educational Concepts, and Large Urology Group Practice Association; Grant funding - NCI Cancer Center Support grant (P30CA006973), UM1 grant (UM1CA186691), and R01 grant (R01CA235681)

**JJM:** Consultant - Merck, AstraZeneca, Janssen, BMS, UroGen, Prokarium, Imvax, Pfizer, Seagen/Astellas, Ferring, CG Oncology, Calibr, Immunity Bio, Protara, Photocure

**SD**: Consultant – AstraZeneca, BMS, CG Oncology, enGene, Ferring, ImmunityBio, Johnson&Johnson, Eli Lilly, Pacific Edge, Pfizer, Photocure, Protara, UroGen, Vesica Health.

**TWF**: Stock – Aurora Oncology; Research Funding – Novartis, Bavarian Nordic, Dendreon, GTx, Janssen, Medivation, Sanofi, Pfizer, BMS, Roche/Genentech, Exelixis, Aragon Pharmaceuticals, SOTIO, Tokai Pharmaceuticals, AstraZeneca/MedImmune, Lilly, Astellas Pharma, Agensys, Seagen, La Roche-Posay, Merck, Myovant Sciences, Criterium; Patents – early-stage bladder cancer treatment and detection (neither commercialized or in active clinical development).

**SPL**: Clinical Trials - Aura Biosciences, Ferring, JBL, Genentech, Merck, Surge Therapeutics, Tyra Biosciences, Vaxilion, Viventia; Consultant – Aura Biocience, BMS, Ferring, ImmunityBio, Incyte, Gilead, Pfizer/EMD Sernoo, Protara, Surge Therapeutics, Tyra Biosciences, UroGen, Vaxiion, Verity; Patent – TCGA classifier; Honoraria – Grand Rounds Urology, UroToday.

**GDG:** Artera AI, MyCareGorithm

**WJS:** Consultant - Urogen

**DJM**: Research support - Convergent Genomics, Ferring; Consultant - CG Oncology, Owkin

**PES**: No financial COI; leadership role on NCCN bladder and penile cancer panel (vice-chair) and board of directors as well as steering committee.

**FZ, YH, PD:** Employment – Predicine

**SJ:** Leadership/Employment - Predicine

**JL, PM, KR, YP, LC, AP, LZ, AY, XW, HX, MAP, SMG** – All declare no COI

## Abbreviations

AJCC: American Joint Committee on Cancer
AUC: area under the curve
BCG: Bacillus Calmette- Guérin
CIS: carcinoma in situ
CHIP: clonal hematopoiesis of indeterminate potential
CNB: copy number burden
CNV: copy number variation
G-score: genomic score
HG: high-grade
HR: hormone receptor
IQR: interquartile range
iTURBT: index transurethral resection of bladder tumor
LP-WGS: low-pass whole genome sequencing
MIBC: muscle-invasive bladder cancer
MRD: minimal residual disease
NGS: next-generation sequencing
NMIBC: non-muscle-invasive bladder cancer
PDD: photodynamic diagnosis
PBMC: peripheral blood mononuclear cell
SCC: squamous cell carcinoma
SOC: standard of care
TF: tumor fraction
TURBT: transurethral resection of bladder tumor
utDNA: urinary tumor DNA
VAF: variant allele frequency
WES: whole-exome sequencing
WGS: whole-genome sequencing

## Data Availability

All data produced in the present study are available upon reasonable request to the authors

---

**Supplemental Figure 1.**
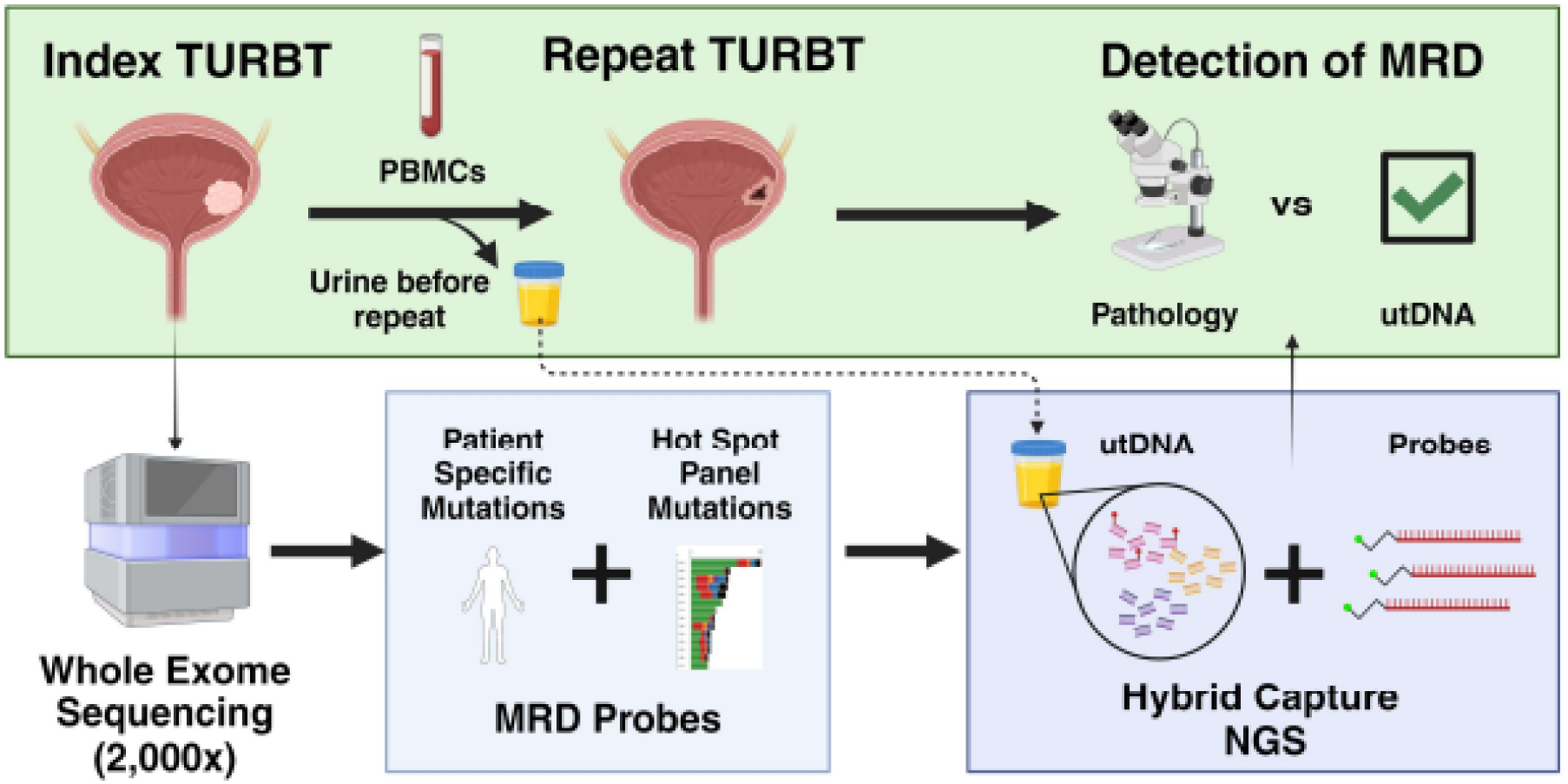
Study schema. Whole-exome sequencing was performed on index transurethral resection of bladder tumor (TURBT) tissue samples and matched peripheral blood mononuclear cells (PBMCs) as germline controls to generate patient-specific mutational probes for minimal residual disease (MRD) detection. In parallel, low pass whole genome sequencing was performed on tumor samples to estimate the sample-level copy number burden (CNB). Urine samples were collected prior to scheduled repeat TURBT (reTURBT) and analyzed for the presence of both patient-specific and panel-based mutations, as well as CNB. Urine MRD results were compared with reTURBT pathology findings, which served as the reference standard.

**Supplemental Figure 2.**
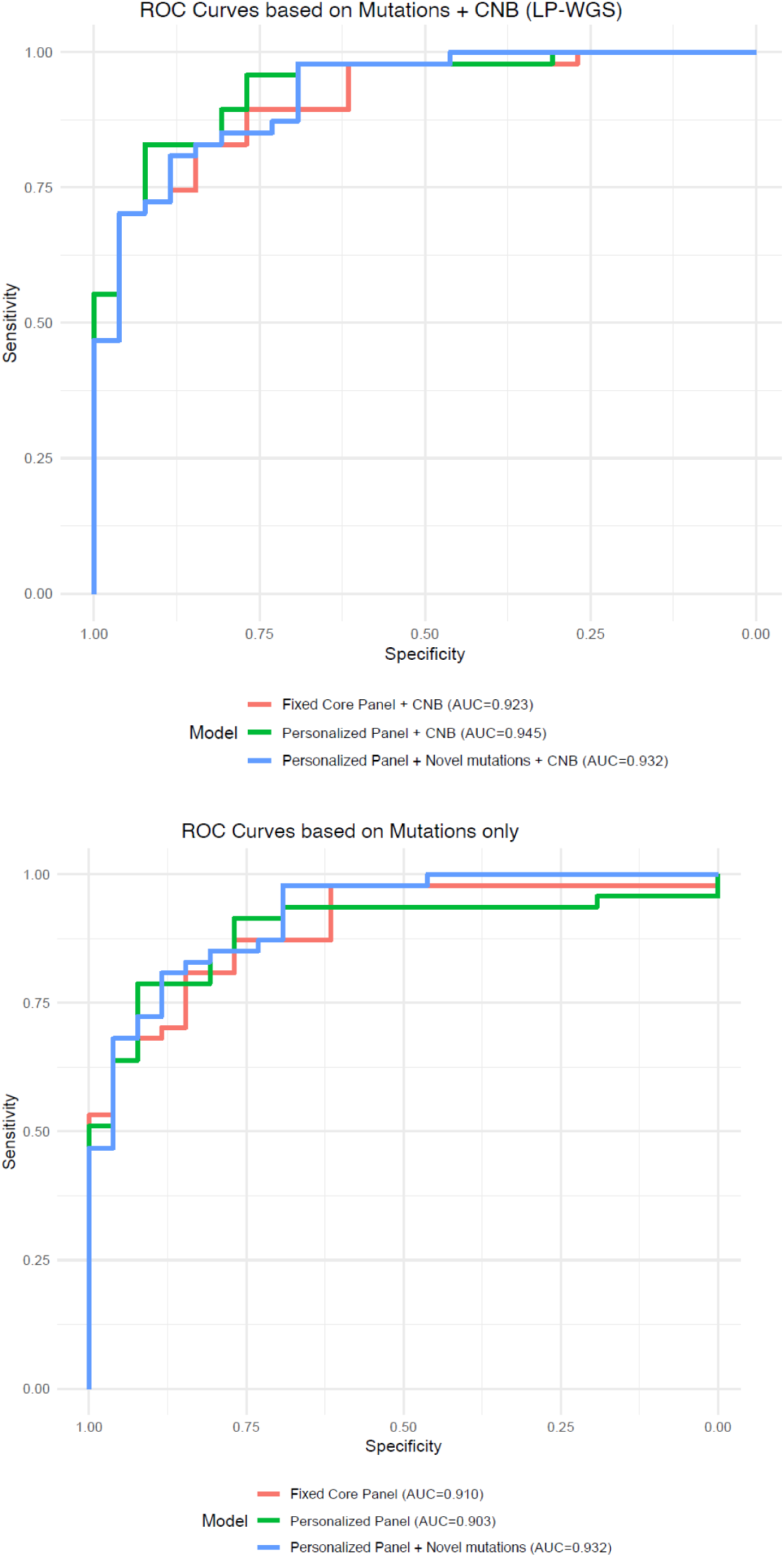
Area under receiver operating characteristic (AUROC) curves comparing tumor fraction (TF) with personalized versus panel mutations, and combinations of TF with or without copy number burden (CNB) score. A) Comparison of AUC based on TF from fixed core panel, personalized panel, or both, with the addition of CNB score. B) Comparison of AUC with TF from fixed core panel, personalized panel, or both, but without CNB score.

**Supplemental Table 1.**
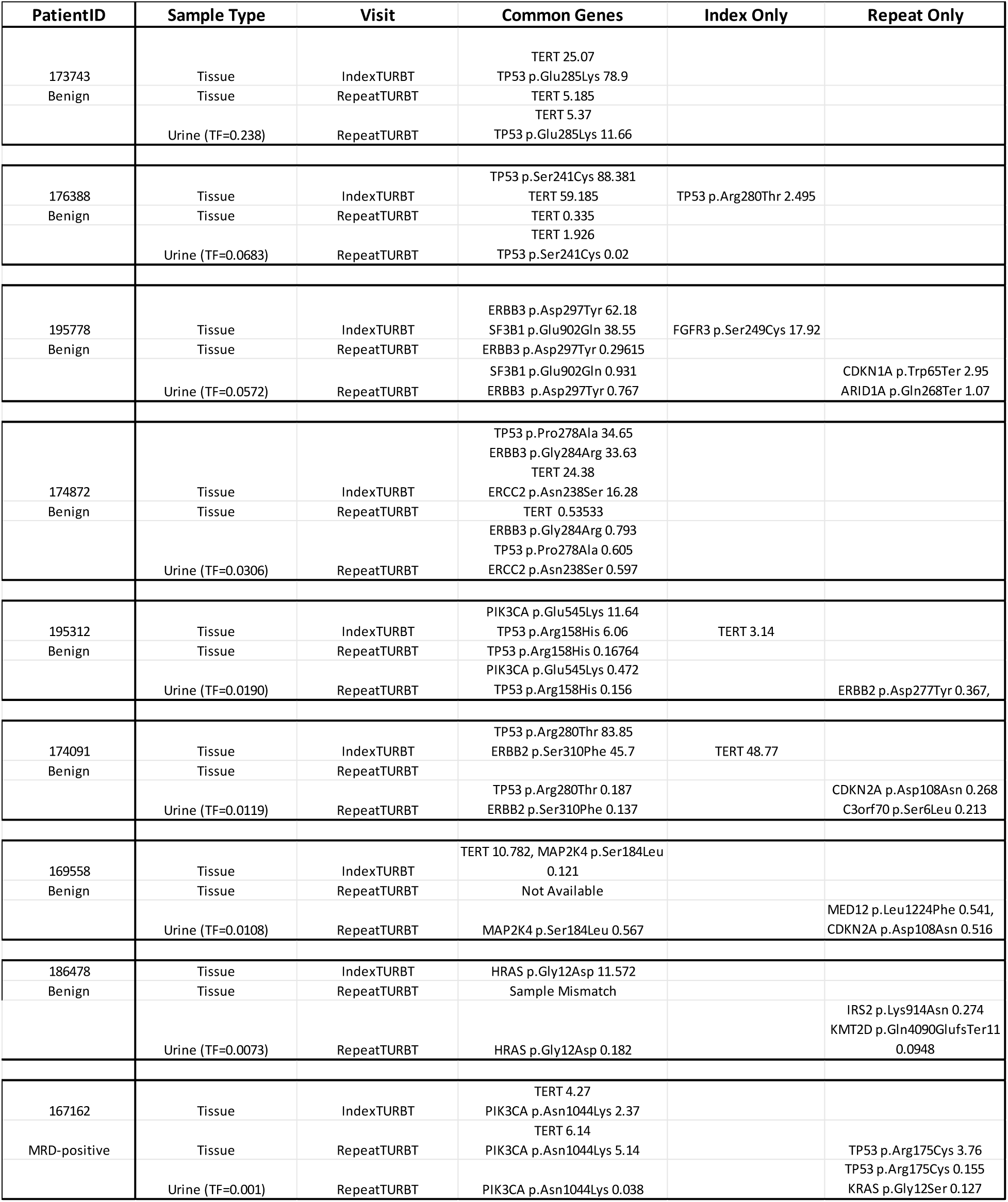
Individual gene variants detected in each tissue type for patients with discrepant results between urinary tumor DNA (utDNA) assay and repeat transurethral resection of bladder tumor (reTURBT) pathology.

